# Premature Coronary Artery Disease Related Mortality in the United States: Regional, Gender, and Racial Disparities – Insights from the CDC WONDER Database (1999–2023)

**DOI:** 10.64898/2025.12.16.25342435

**Authors:** Iftikhar Ali Ch, Soban Ali Qasim, Hussnain Zafar, Shaiza Maryam, Iftikhar Khan, Muhammad Qasim, Saif Ur Rahman, Ankur Kalra, Khurram Nasir

**Author notes:** **Corresponding Author:** Iftikhar Ali Ch MD, SSM Health St Anthony Hospital, Oklahoma City, OK, USA.

## Abstract

**Background:** Premature coronary artery disease (PCAD) continues to impose a disproportionate burden on younger adults in the United States, yet recent patterns across sex, region, race, and urbanization remain poorly defined.

**Methods:** Using CDC WONDER data from 1999–2023, we examined PCAD-related age-adjusted mortality rates (AAMR) for males <45 years and females <55 years, stratified by region, race/ethnicity, and urbanization. Temporal trends were assessed using Joinpoint regression to estimate annual percent change (APC) and average annual percent change (AAPC).

**Results:** The combined AAMR for PCAD in the U.S. is approximately 8.5 deaths per 100,000 population. Both sexes demonstrated overall declines in PCAD mortality since 1999 (AAPC males −0.84%; females −1.07%), interrupted by a transient rise during 2018–2021 (APC = 7.85%; 95% CI 5.41–9.32), followed by a sharp post-pandemic decline (APC = −7.39%; 95% CI −10.64 to −4.53). Females consistently exhibited higher mean AAMRs than males (8.74 vs. 8.34; p<0.00001). Regional analyses showed that mortality rates were highest in the South (males 9.86; females 10.96) and Midwest (8.97; 9.36), with intermediate rates in the Northeast (males 7.09, females 6.77), and the lowest rates in the West (6.35; 6.16). Non-metropolitan residents carried a 1.5–1.7-fold greater mortality burden than metropolitan populations (males 12.20 vs. 7.74; females 13.31 vs. 8.06). Black/African Americans had the highest rates (males: 12.17, females: 15.98), followed by American Indian/Alaska Natives (8.49, 9.02) and Whites (8.0, 8.0), while Asian/Pacific Islanders had the lowest (about 2.4–4.3).

**Conclusions:** National PCAD mortality has decreased, but disparities persist and are growing by region, race, sex, and urbanization. Concentration in the Southern and Midwestern states and among certain races highlights the need for further research using modern molecular methods and improved health care resources.

**Graphical Abstract:** 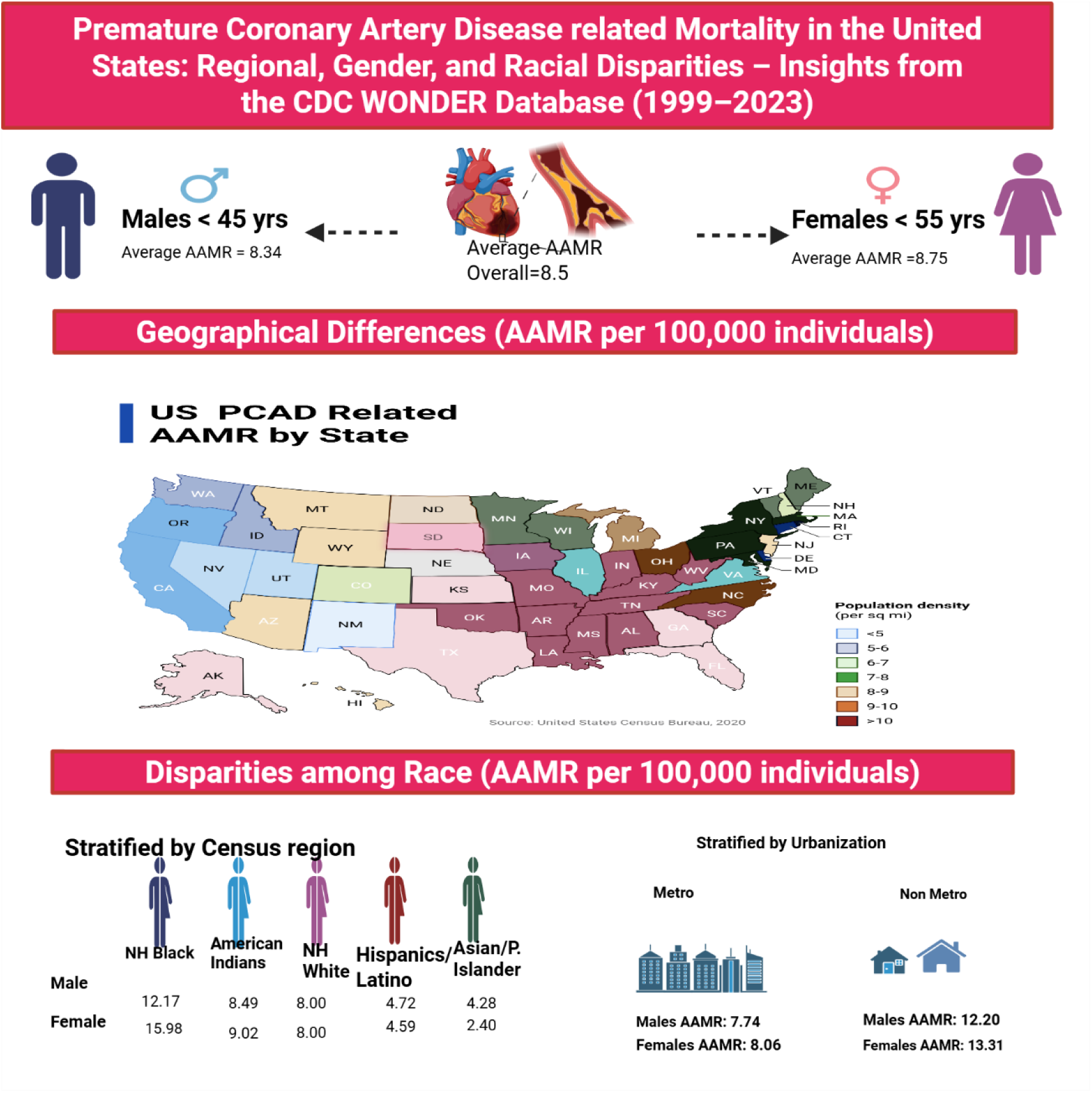

**Clinical Perspective:** *What is New?:*

- The combined AAMR for PCAD in the U.S. is approximately 8.5 deaths per 100,000 population.
- Premature coronary artery disease–related mortality rates are higher in the Southern and Midwestern states, among non-Hispanic Black, Native American, and non-metro populations.

*What are the Clinical Implications?:*

- The geographic and demographic clustering of cases strongly suggest contributions from underlying genetic, environmental, and dietary factors that warrant further investigation.
- Early identification of at-risk individuals through precision-medicine approaches and the implementation of targeted, evidence-based preventive strategies could mitigate these regional and racial disparities.

## Introduction

Recent studies also show that cardiovascular disease (CVD) is becoming more prevalent among younger populations around the world ^1^. In spite of the advances in prevention and treatment, the global rates of CVD are expected to increase by about 90 per cent in the next 25 years ^2^. Premature CAD has been defined in terms of age with different parameters applied to each gender, that is, coronary artery disease at a young age (CAD). A few studies use the Framingham criteria, where men are younger than 55 years and women are younger than 65 years ^3,4^, and have found myocardial infarction (MI) mortality in the middle-aged (45-65 years) and the younger (<45 years) groups. Other studies use age limits that are more restrictive in order to determine younger cohorts. The common understanding of premature CAD is male below 45 years and female below 55 years ^5^. Multiple studies consistently find an increase in CVD incidence among younger individuals internationally ^6,7,8^.

The Global Burden of Disease study reported that between 1999 and 2019, cases of CVD before age 50 rose by 25%, resulting in more than 1.2 million deaths ^1^. Although population-adjusted incidence rates have decreased, the overall impact remains substantial, especially among adults aged 40–49 years, where mortality rates nearly double between the 40–44 and 45–49 groups, with about 60 deaths per 100,000 individuals ^1^. Assessment of premature mortality due to acute MI may inform health care policy, though a comprehensive evaluation of premature CVD should include deaths related to both ischemic heart disease and MI, including their complications. Dani et al. documented higher age-adjusted mortality rates (AAMR) due to MI and its complications in the middle-aged group (34.9 [95% CI, 34.8–35.0]) compared with the younger group (2.5 [95% CI, 2.4–2.6]) (4). The burden appears lower among younger populations, but this may underestimate the actual disease burden for several reasons: (a) only MI and MI complication ICD codes were included, excluding ischemic heart disease codes 123–125; (b) gender and racial disparities may be understated; and (c) improved survival rates in younger people could give a misleading impression of lower burden of disease relative to older groups. Accordingly, an analysis was conducted of AAMR for premature CAD using age criteria of under 45 years for males and under 55 years for females and incorporating ICD codes 120–125. While mortality from MI or ischemic heart disease in younger populations does not provide a complete estimate of disease burden, it offers a more precise evaluation of premature disease impact on survival within this age group.

## Methods

### Study Setting and Population

In this descriptive study, we utilized mortality data from the Centers for Disease Control and Prevention (CDC) Wide-Ranging Online Data for Epidemiologic Research (WONDER) Multiple Cause-of-Death database ^9^. We analyzed mortality that was experienced in the United States between 1999 and 2023. Premature coronary artery disease (CAD) mortality was categorized as per the age based-related criteria where premature disease was categorized as in men below 45 years and women below 55 years. The International Classification of Diseases, Tenth Revision I20-I25 was used to identify cases of ischemic heart diseases, which include acute myocardial infarction, chronic ischemic heart disease, and associated coronary pathology ^10^. Multiple Cause-of-Death Public Use files were searched to get death certificates that reported a CAD-related cause of death (I20-I25) being the underlying cause of death among those who met the age requirement to have premature CAD. All the information was de-identified, publicly accessible, and was not enclosed with individual identifiers. Thus, there was no need to have institutional review board approval. The reporting was done in compliance with STROBE (Strengthening the Reporting of Observational Studies in Epidemiology guidelines ^11^.

### Data Abstraction

We abstracted the number of deaths attributable to coronary artery disease (CAD) from 1999 to 2023, along with the corresponding annual population estimates. CAD-related deaths were identified using ICD-10 codes I20–I25. For this study, we focused on premature CAD mortality, defined as occurring among male decedents aged <45 years and female decedents aged <55 years. From each death certificate, we extracted demographic variables including age, sex, race/ethnicity, and geographic location at the state and county levels. Race and ethnicity were classified as non-Hispanic (NH) White, NH Black, Hispanic, NH American Indian/Alaska Native, and Asian/Pacific Islander. Counties were categorized according to the National Center for Health Statistics (NCHS) 2013 Urban–Rural Classification Scheme and grouped into large/medium metropolitan, small metropolitan, and rural (micropolitan and noncore) counties.

### Statistical Analysis

We calculated age-adjusted mortality rates (AAMRs) per 100,000 population for premature CAD by sex, race/ethnicity, census region, and urbanization level from 1999 to 2023. Age-adjustment was performed using the 2000 U.S. standard population as the reference ^12^. Only age-adjusted mortality rates available directly from the CDC WONDER database were used. Temporal trends in premature CAD mortality were evaluated using the Joinpoint Regression Program (Version 5.2.0.0, National Cancer Institute) to estimate the annual percent change (APC) and average annual percent change (AAPC) with corresponding 95% confidence intervals ^13^. The Joinpoint model identifies statistically significant shifts in mortality trends over time by fitting log-linear regression segments. A trend was considered statistically significant if the APC or AAPC was different from zero, based on a two-sided p value <0.05.

### Results

Between 1999 and 2023, the total number of deaths was 183,384 among males and 259,880 among females. The age-adjusted mortality rate (AAMR) was slightly higher in females (8.74) compared with males (8.34). Figure 1 presents data on PCAD-related mortality.

**Fig 1:**
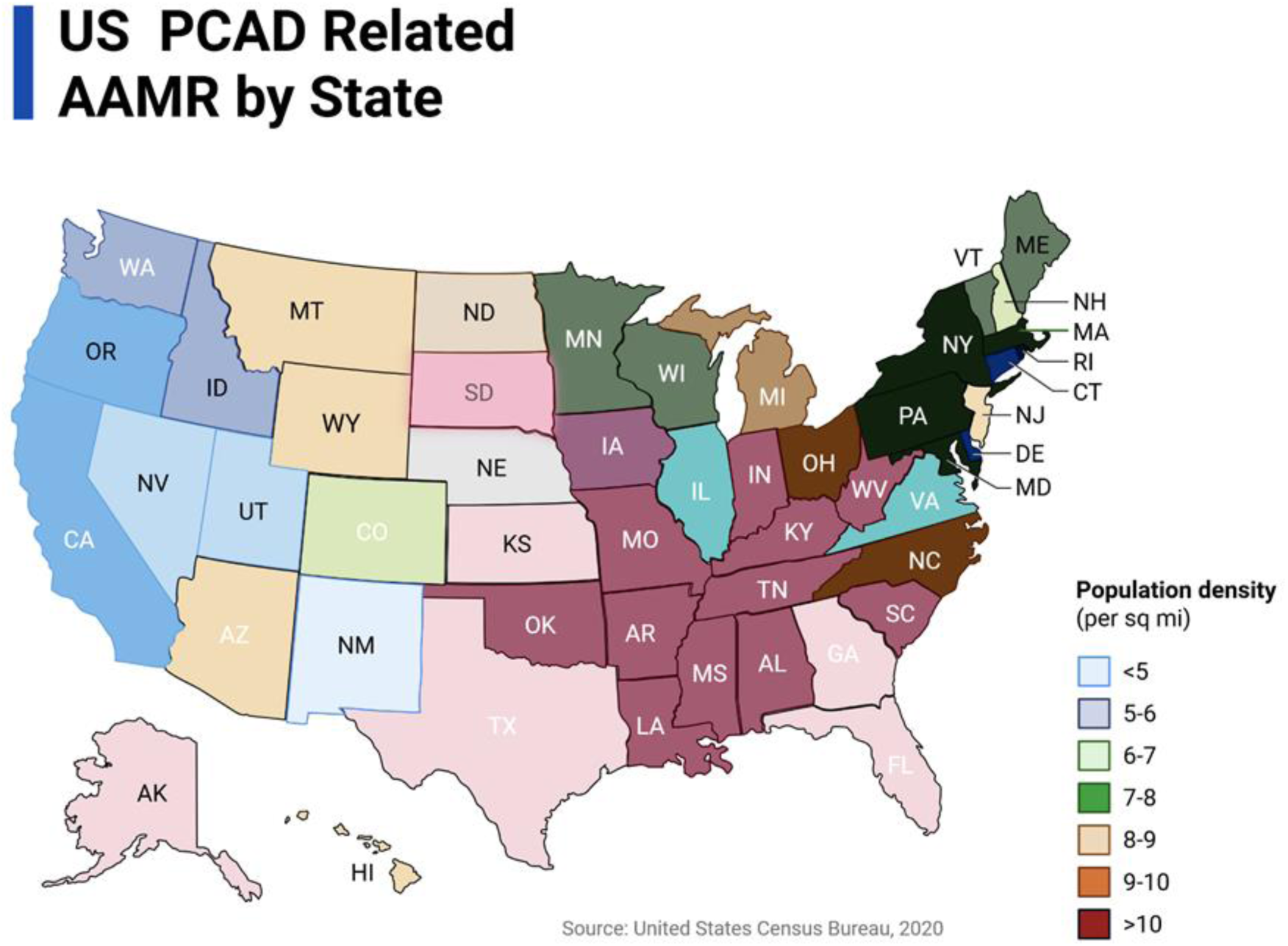
Geographic distribution of age-adjusted mortality rates (AAMRs) for premature coronary artery disease across U.S. states (1999-2023).

### Trends in PCAD Mortality by Gender

Between 1999 and 2023, both males under 45 years and females under 55 years exhibited an overall declining trend, though with notable fluctuations over time. Females had higher mean AAMR of (8.74; 95% CI 8.58-8.92) compared with males (8.34; 95% CI 8.16-8.52). The combined analysis showed a modest rise from 1999 to 2003 (APC = 1.35%; 95% CI 0.12–3.60), followed by a sharp decline between 2003 and 2009 (APC = −2.96%; 95% CI −4.67 to −2.23) and a slower continued decrease from 2009 to 2018 (APC = −1.65%; 95% CI −2.33 to −0.24). This pattern reversed between 2018 and 2021 with a pronounced increase (APC = 7.85%; 95% CI 5.41–9.32), before dropping steeply again from 2021 to 2023 (APC = −7.39%; 95% CI −10.64 to −4.53). Overall, the average annual percent change (AAPC) indicated sustained long-term declines for both males (−0.84%; 95% CI −1.11 to −0.62) and females (−1.07%; 95% CI −1.45 to −0.86), despite short-term upswings in recent years (Figure 2 male and female overall); Tables S1, S2, S4).

**Fig 2:**
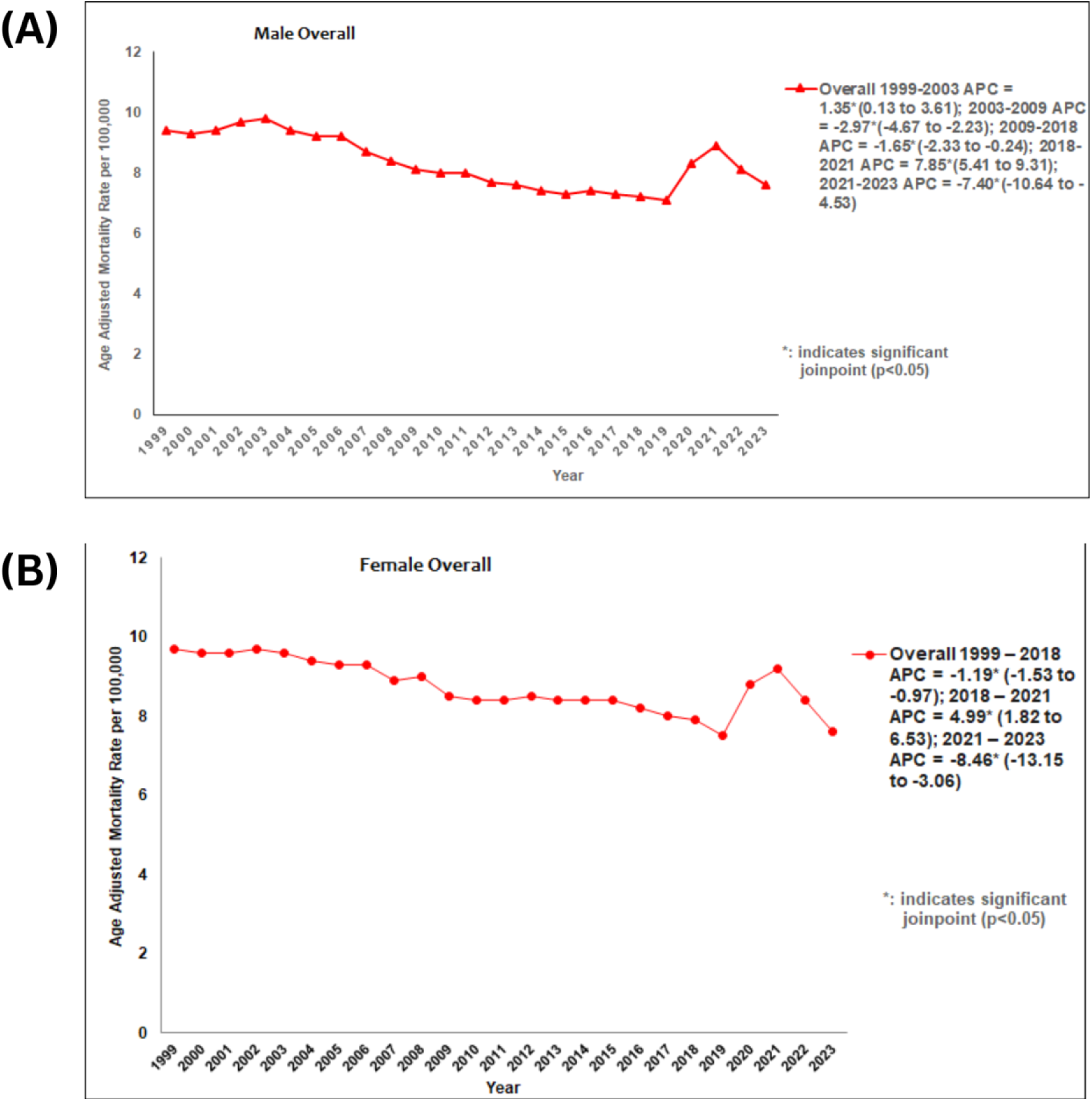
Trends in premature coronary artery disease mortality in the United States from 1999 to 2023: (A) males aged <45 years and (B) females aged <55 years.

**Fig 3:**
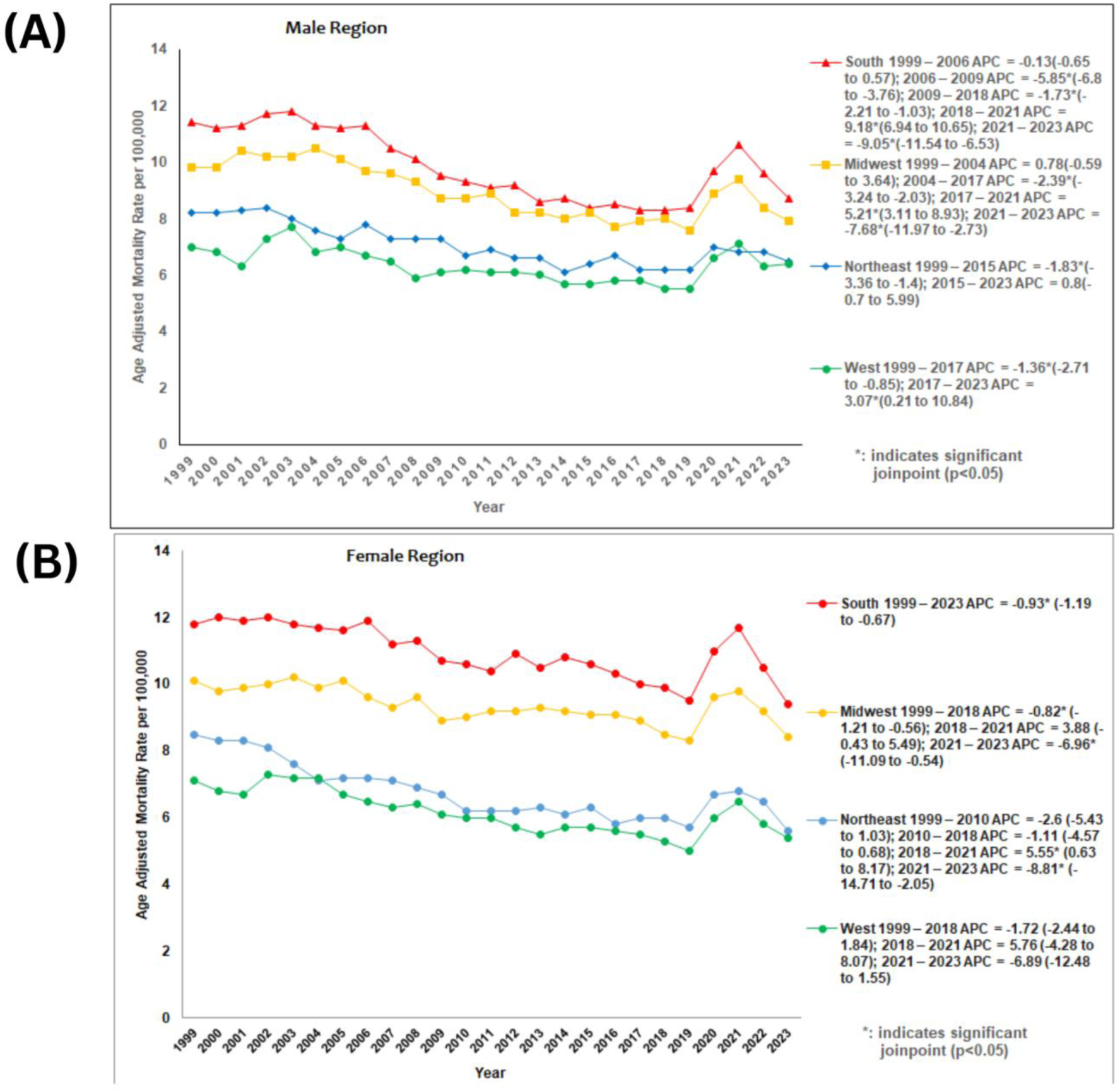
Trends in premature coronary artery disease mortality in the United States from 1999 to 2023, stratified by Region: (A) males aged <45 years and (B) females aged <55 years.

### Census/Regional disparities

Regional analysis revealed notable geographic and sex-specific disparities in AAMR between 1999 and 2023. The Southern region exhibited the highest AAMRs for both males (9.87; 95% CI 9.52-10.20) and females (10.96; 95% CI 10.66-11.26). The Midwest followed with elevated rates (males 8.97; 95% CI 8.54-9.42), females 9.37; 95% CI 8.98-9.74), whereas the Northeast showed intermediate mortality (males 7.10; 95% CI 6.68-7.52), females 6.78; 95% CI 6.44-7.12). The West consistently demonstrated the lowest AAMRs (males 6.36; 95% CI 6.01-6.70), females 6.16; 95% CI 5.85-6.45), suggesting more favorable cardiovascular outcomes. Overall, these data reveal a persistent South-to-West gradient, with mortality highest in the South and lowest in the West, and females exhibiting slightly higher rates than males in the South and Midwest. Across regions and in both genders, long-term declines were punctuated by brief surges between 2018 and 2021 followed by sharp reversals (to see trend refer to supplement S1,S2,S6,S9, Figure3).

### Urbanization

Major differences were noted on the basis of urbanization. The non-metropolitan areas had significantly higher burden of mortality, and the non-metro males had an average PCAD AAMR of (12.21; 95% CI 11.57-12.84), as compared to (7.74; 95% CI 7.54-7.94) in metro males, and (13.32; 95% CI 12.77-13.88) in non-metro females. The results show that the age-adjusted mortality rates are almost 1.5 to 1.7 times higher among the rural population, which points to a consistent and severe urban-rural disparity in cardiovascular mortality in both sexes. Early plateau followed by a steep decrease in metro males (APC = 7.53%; 95% CI 2.77-10.73; p < 1×10-6) and a steep rise in non-metro males (APC = 1.43%; 95% CI −2.36- −1.09; p = 0.001) followed by a significant decrease thereafter. Metro females demonstrated a significant long-term decrease (APC = −1.64%; 95% CI −2.06- −1.43); p = 0.01) with a minor, non-significant uptick after 2018, whereas non-metro females showed a modest but significant overall rise (APC = 0.89%; 95% CI 0.59-1.22; p < 1×10⁻⁶). Collectively, these findings underscore persistent sex-based and urban–rural disparities, with non-metro patients (women more than men) experiencing the highest and most concerning mortality trends (Fig 4; Table S1, S2, S7, S10)

**Fig 4:**
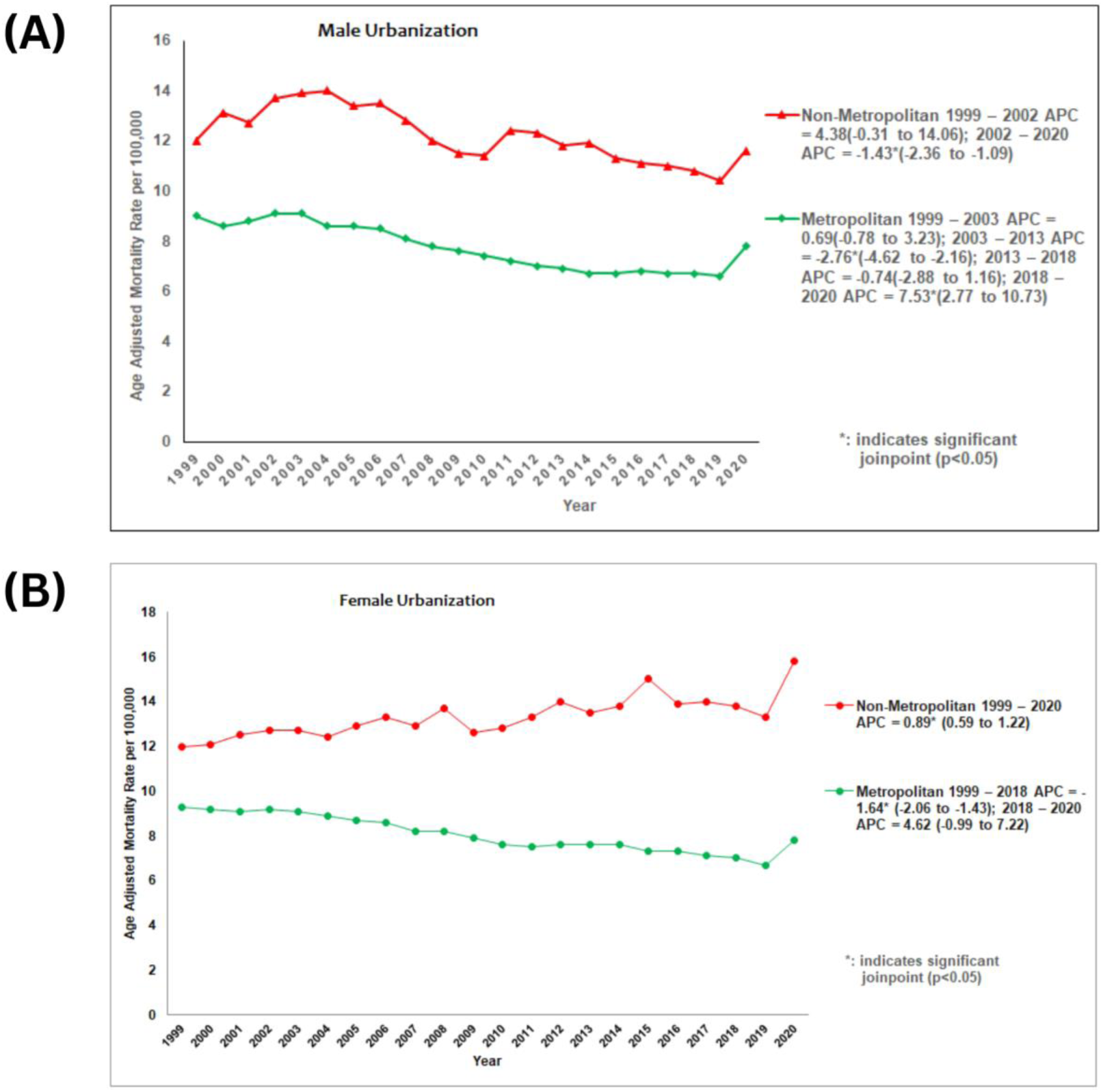
Trends in premature coronary artery disease mortality in the United States from 1999 to 2023, stratified by Urbanization: (A) males aged <45 years and (B) females aged <55 years.

**Fig 5:**
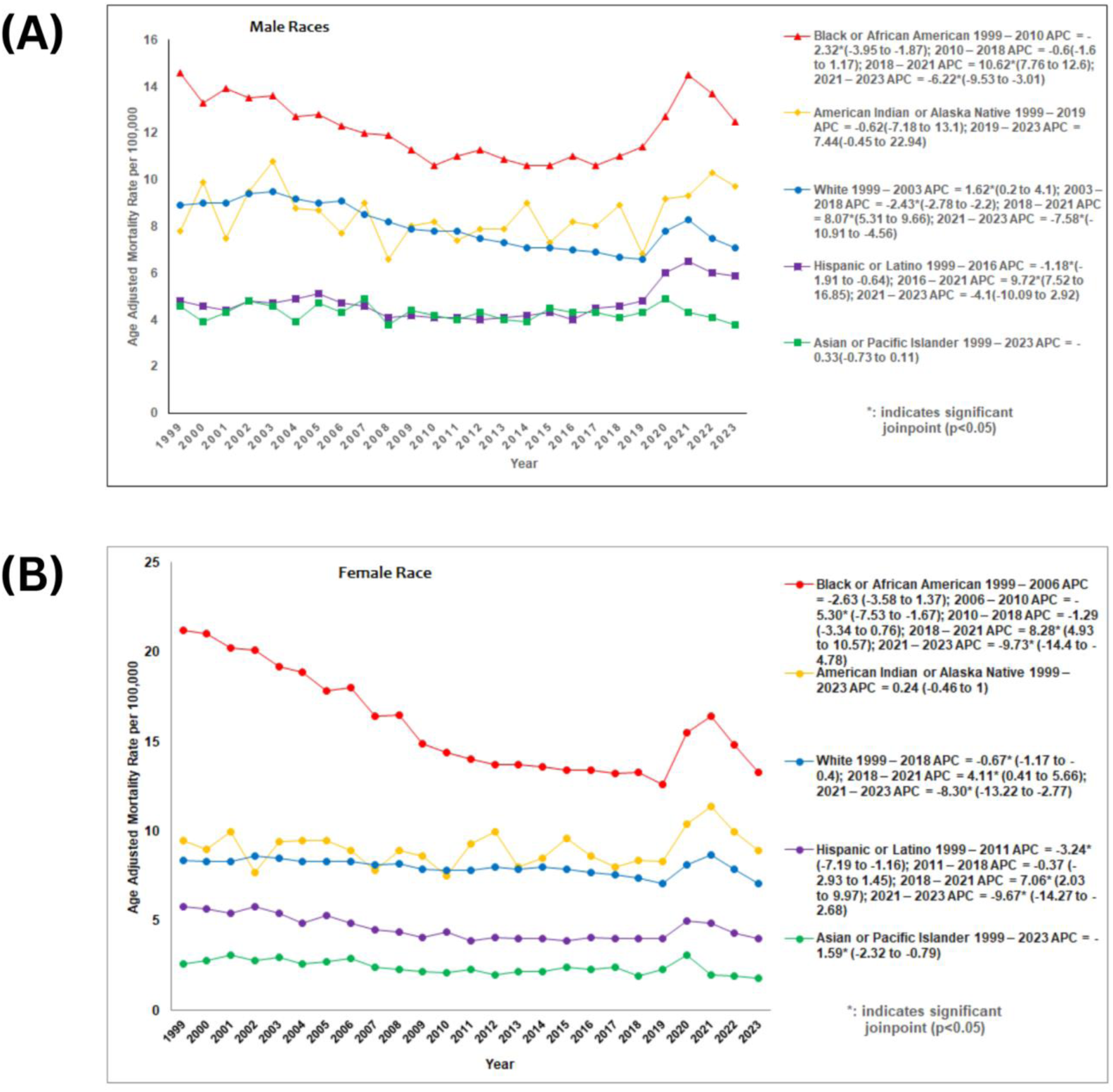
Trends in premature coronary artery disease mortality in the United States from 1999 to 2023, stratified by Race: (A) males aged <45 years and (B) females aged <55 years.

### Race

From 1999–2023, PCAD–related mortality remained highest among Black/African Americans (males 12.17; 95% CI 11.54-12.80, females 15.98; 95% CI 15.36-16.59 per 100,000), followed by American Indian/Alaska Natives (males 8.50; 95% CI 6.88-10.28, females 9.03; 95% CI 7.46-10.61), Whites (males 8.00; 95% CI 7.80-8.22, females 8.01; 95% CI 7.83-8.19), Hispanics (males 4.72; 95% CI 4.37-5.07, females 4.59; 95% CI 4.24-4.92), and Asian/Pacific Islanders (males 4.29; 95% CI 3.73-4.83, females 2.41; 95% CI 2.04-2.78). Both genders showed early declines, sharp increases around 2018–2021, and subsequent drops thereafter. Black males (APC = −2.32%; 95% CI −3.95- −1.87 to 2010; +10.62%; 95% CI 7.76-12.6 in 2018–2021; −6.22%; 95% CI −9.53- −3.01 after 2021) and females (−5.30%; 95% CI −7.53- −1.67 to 2010; +8.28%; 95% CI 4.93-10.57 in 2018–2021; −9.73%; 95% CI −14.4- −4.78 after 2021) exhibited the largest fluctuations. Whites and Hispanics followed similar but less pronounced patterns, while American Indian/Alaska Native and Asian/Pacific Islander groups showed smaller or nonsignificant changes. Overall, racial disparities persisted, with Black populations consistently experiencing the highest and most volatile mortality trends (Fig 4; Table S1, S2, S5, S8)

## Discussion

### Gender disparities

In this nationwide analysis of PCAD mortality, defined as deaths occurring in men younger than 45 years and women younger than 55 years, we observed persistent differences by sex, region, and urbanization, though not always in the hypothesized direction or magnitude. Overall, females had a slightly higher age-adjusted mortality rate (AAMR) than males (8.74 vs. 8.34), consistent with prior evidence suggesting younger women face a disproportionate risk once CAD develops. Women under 55 admitted with acute coronary syndromes had a higher risk of in-hospital death (HR 4.1), 30-day major adverse events (HR 2.1), and 5-year mortality (HR 1.96) compared with men, pronounced in the context of STEMI and among women with significant comorbidities such as diabetes and hypertension ^14^. Unique risk factors in women, including early menopause, higher prevalence of diabetes and hypertension, and atypical symptom presentation, contribute to poorer outcomes ^15, 16, 17^ and receive poorer secondary preventive care (16). Large international cohort studies suggest that, after adjustment for comorbidities and risk factors, long-term survival may be similar or slightly better in women than men with established CAD, but this advantage is less pronounced in younger women and those with STEMI ^18, 19^. In summary, young women with premature CAD have a survival disadvantage compared with men, driven by both biological and healthcare disparities, and this gap is most evident in the youngest age groups and those with acute presentations.

### Overall trend

Between 1999 and the end of the 2010s, both genders showed a consistent decline in mortality. Such continuous declines in cardiovascular deaths can be probably explained by the progress in cardiac medical care, pharmacotherapy, interventional approaches, the increased access to coronary operations, and the favorable changes in the risk factors of the population ^20, 21,22,23,24^. Nevertheless, since around 2011, mortality rates (AAMR) have been decreasing at a standstill in most of the United States. This slowdown is multivariate, which can be caused by the rise in hypertension, smoking, diabetes, and obesity ^21, 25, 26, 27^. There was a significant recovery between 2018 and 2021 (APC = 4.98% in women, 95% CI 1.82-6.53; APC = 7.85% in men, 95% CI 5.41-9.32), after this stagnation. This tendency in these years is probably the result of both the cumulative effects of an already slowing down decrease and the significant, acute influence of the COVID-19 pandemic.

### Pandemic effect

The COVID-19 pandemic adversely impacted the cardiovascular mortality trends. It is common for adults hospitalized with COVID-19 to have acute cardiac events and those who recover are at an increased risk of cardiovascular events ^28, 29, 30^. Moreover, patient hesitancy to seek care and disrupted healthcare systems may have delayed detection and treatment of cardiovascular diseases and risk factors ^31, 32^. Disruptions due to COVID-19 hampered efforts to engage in healthy behaviors leading to increased obesity and hypertension^33, 34^. Additionally, hospitals were overburdened by the pandemic, leading to overflowing ICU, emergency departments and shortage of beds, contributing to decreased critical care capacity ^35,36^. This surge in mortality was followed by sharp declines through 2023 (APC = −8.45% for females, 95% CI −13.15 to −3.05; APC = −7.39% for males, 95% CI −10.64 to −4.53) This strong decline from 2021 through 2023 is temporally consistent with decreases in COVID-19 mortality, partial recovery of health-care infrastructure and COVID-19 vaccination rollout ^37^. Use of tele-medicine to manage risk factors and chronic cardiovascular diseases also likely improved outcomes ^38^. Lastly, newer and more efficacious treatments like SGLT-2 inhibitors and GLP-1 receptor agonists, along with advancements in intravascular imaging led to a decrease in risk factors associated with cardiovascular disease and reduced cardiovascular mortality ^39,40,41^.

### Regional disparities

Regional analyses revealed that the South consistently bore the highest AAMR for both sexes (females: 10.96, males: 9.86), while the West had the lowest (females: 6.16, males: 6.35). This pattern echoes the entrenched “Southern heart-disease belt” described in CDC reports ^42^. Western and Northeastern females experienced declines that were non-significant but still kept their baseline mortality far lower (e.g., Western females AAMR 6.16 vs. Southern females 10.96). Even when downward trends are observed, the South remains an outlier with persistently higher absolute mortality. These excesses reflect deep structural disadvantages unique to the South, including scarce specialty cardiology resources, limited healthcare infrastructure, and low insurance coverage, higher burden of CV risk factors, longer travel distances to tertiary centers, and pervasive socioeconomic deprivation ^43, 44,45,46,47^. A study showed a causal relationship between higher altitude and lower cardiovascular mortality, which could account for the lower cardiovascular mortality in the Western and Southwestern states^48^. Additionally, local factors may influence effect modification, contributing to disparities, as observed risk associations vary across different regions of the country ^49^. In summary, the Southern United States not only bears the greatest burden of PCAD but is also the region with the least capacity to address this challenge. It highlights the urgent necessity of the specific investments, region-specific prevention plans, and increasing healthcare accessibility to reduce this loop of disproportional deaths. The prevalence of CAD, particularly the premature cases in the South, has always been high, indicating that there might be other factors that contribute to the disease and make the region disadvantaged in terms of healthcare infrastructure. It is worth noting that the differences between the South and other areas like the Northeast, and West cannot be explained entirely by the differences in healthcare. The future studies ought to adopt integrative omics methods such as genomics, proteomics and metabolomics to determine region-specific biological and metabolic predictors of cardiovascular risk. The peculiarities of diet, environmental contacts, and genetic ancestry profiles of the populations in these states can all lead to their disproportionate disease burden.

### Urbanization effect

The patterns of urbanization were clear excess mortality in the nonmetropolitan regions with non-metro females (AAMR = 13.31) and males (AAMR = 12.20) exceeding that in the metropolitan regions (females: 8.06, males: 7.74). Non-metro females showed a small significant increase between 1999-2020, whereas the metro females showed a significant decrease between 1999-2018. A borderline rise was observed in non-metro males between 1999-2002, which was followed by a drastic drop in the years 2002-2020. These results are in line with CDC and American Heart Association reports that recorded rural health disparities in cardiovascular health, which are associated with inadequate healthcare facilities, increased travel distance to tertiary health centers, decreased insurance coverage, and increased cases of smoking, obesity, and diabetes ^50,51,52^. The increased mortality rate among non-metro females is attributed to various factors such as atypical symptom presentation ^53, 54^, diagnostic problems and under treatment (^55, 56^), and a rise in cardiovascular risk factors such as hypertensive disorders in pregnancy which increase lifetime risks of cardiovascular mortality ^57, 58^. Such concerns are reported in the past by Dani and others Dani SS, 2022 Jan.

### Racial disparities

Racial and ethnic disparities, while not directly detailed in the primary results, are supported by a robust literature base indicating higher cardiovascular mortality among non-Hispanic Black populations and American Indian and Alaska Native groups compared with non-Hispanic Whites ^59,60,61^. The reasons for this are multi-fold. One is genetic predisposition. Studies have reported that Black individuals tend to develop hypertension at a comparatively younger age and with greater severity ^62^, partly due to increased salt sensitivity ^63^. Additionally Black individuals suffer from more comorbidities like diabetes, chronic kidney disease, obesity, hypertension, and coronary artery disease ^64,65^. Next are social and structural inequalities. It is reported by several studies that hospitals serving major African American and American Indian populations receive lower funding than other hospitals which results into poor health quality ^66,67^. An important factor is healthcare insurance. There is a higher number of uninsured individuals or Medicaid users in the Black population (5–10 %, 16–23 %) compared with the White population (2–5%, 6–11 %) ^68,69^. Other sociocultural factors like psychosocial stressors, racism in physician-patient interactions, decision-making, and utilization of healthcare services also exist ^68^. Lastly, poverty, residential segregation, and food insecurity likely contribute to these disparities ^70,71,72^.

These results bring out a paradox on progress in premature CAD mortality. The long-term decreases indicate the achievements in the prevention, acute management, and secondary prevention ^3,1^, but the reversal of 2018-2021^73^, the Southern concentration ^74^, the long-standing rural disadvantage ^75^, and the racial disparities ^76^ demonstrate that some problems still exist. The excess mortality burden in the South (20-30 percent) highlights the necessity of interventions, including the expansion of telecardiology or the screening of risk factors ^43^. A study of CDC WONDER data 2025 revealed that sudden cardiac death (SCD) mortality had decreased by 23% since 2018 ( AACP = [?]1.20%), then rose by 6.93% between 2018-2020, due to pandemic-related disruptions and socioeconomic barriers, especially those in the South ^77^. We have a CAD-specific reversal (5-8% increase) that is slightly weaker but consistent, and similar rural-urban results are reported in a 2021 CDC report that reported 21% higher death rates of heart disease in rural areas ^35^. The South has increased racial inequalities, and Black and Hispanic individuals are 1.9-2.4 times more likely to die of CVD due to systemic inequalities such as poorer insurance coverage ^54^. This interaction establishes a structural context where younger adults will be susceptible to premature CAD mortality despite national improvements in cardiovascular medicine and perceived reduced CV risk factor burden. The observed geographical distribution of premature CAD in the Southern and Midwestern United States (especially among the Blacks, American Indians, and Whites) indicates that genetic predisposition, environmental exposures, and localized eating patterns might all contribute to the observed excess burden. These results emphasize the importance of integrative omics-based studies, including genomics, proteomics and metabolomics, to understand new biological pathways and mechanisms of atherosclerosis in young population.

## Limitations

Acknowledged limitations are the low statistical power because of low numbers of premature deaths of CAD, which results in broad confidence intervals that can mask any slight discrepancies ^78^. Possible misclassification of cause of death, alterations in coding, and regional migration, and inability of the ecological design to measure risk factors at the individual level can introduce bias ^79, 80, 81^. There are no 2021-2023 data available on certain subgroups (e.g. the non-metro females), which restricts trend assessments. However, standardized ICD-10 definition, long-term national data, and stratified analyses make our findings stronger.

## Conclusion

To sum up, although the premature CAD mortality has decreased during the last 20 years, there are still severe differences. The South has the highest rate of burden, excess mortality is demonstrated in nonmetropolitan areas, and racial/ethnic disparities exist. The causes of the 2018-2021 reversal (e.g. COVID-19 disruptions, risk factor trends), combining granular socioeconomic and behavioral data, assessing regionally-specific interventions to decrease premature CAD mortality in high-risk groups, especially in Southern and rural communities and targeted should be investigated in future work.

## Nonstandard Abbreviations and Acronyms

AAMR: Age-Adjusted Mortality Rate
AAPC: Average Annual Percent Change
APC: Annual Percent Change
WHO: World Health Organization
WONDER: Wide-ranging Online Data for Epidemiologic Research

## Institutional Review Board Statement

No ethical approval was required for this study design, as all data were obtained from publicly available sources.

## Informed Consent Statement

Not Applicable

## Data Availability Statement

The data supporting the findings of this study are openly available in CDC Wonder at [https://wonder.cdc.gov/].

## Acknowledgments

The authors have no acknowledgments to declare.

## Clinical Trial

Clinical Trial number not applicable

## Consent for publication

Not Applicable

## Ethics approval and consent to participate

Not Applicable

